# Protocol for a systematic review and meta-analysis to assess the association and risk factors for hypertension in people living with HIV (PLHIV), compared with the HIV-uninfected population

**DOI:** 10.1101/2024.11.25.24317915

**Authors:** Mariene G Gomes, Sinara Laurini Rossato

**Affiliations:** Postgraduate Program in Health Sciences, Medical College, Uberlândia, Brazil; School of Collective Health, Federal University of Uberlandia, Uberlândia, Brazil; Laboratory of Research and Extension in Epidemiology (Lapex-Epi), Federal University of Uberlandia, Uberlândia, Brazil; Visiting Scientist in the Department of Nutrition at Harvard T.H. Chann School of Public Health

**Author notes:** Corresponding author: Sinara Laurini Rossato Street Name: Av. João Naves de Ávila, 2121 - Santa Mônica, Uberlândia - MG, City, Postal Code, Country: Uberlândia, 38408-202. Email address of authors: SLR, MGG.

**Keywords:** people living with HIV, hypertension, systematic review, high blood pressure, epidemiology

## Abstract

**Introduction:** The effectiveness of antiretroviral therapy has increased life expectancy among people living with HIV (PLHIV), yet conflicting reports persist regarding the incidence of hypertension in this population. Understanding the key risk factors for hypertension in PLHIV is crucial for informing public health policy. This article presents the protocol for a systematic review and meta-analysis aimed at exploring associations and identifying risk factors for hypertension in PLHIV compared to the HIV-uninfected population. By detailing the methodological approach, this protocol ensures consistency, transparency, and rigour throughout the study’s execution.

**Methods and Analysis:** This protocol outlines the planned steps for conducting the systematic review and meta-analysis following the Preferred Reporting Items for Systematic Review and Meta-Analysis Protocols (PRISMA-P) guidelines. Observational studies addressing the association and the risk factors for hypertension in PLHIV, compared to the HIV-uninfected population among participants aged ≥ 18 years will be included. The searche will be performed across Medline (PubMed), Embase, Web of Science, Global Index Medicus with no restrictions on language and year of publication. Additionally, reference lists of included articles will be reviewed. Search results will be imported into the Covidence software, where duplicates will be removed, and data screening, selection, and extraction will be performed. Methodological quality and potential biases will be assessed using the Newcastle-Ottawa scale (NOAS), Checklist for Analytical Cross-Sectional Studies from the Joana Briggs Institute (JBI), and Grading of Recommendations Assessment, Development, and Evaluation (GRADE). All stages will be completed by three reviewers independently. Data will be systematically analysed and summarised and a meta-analysis will be performed.

**Ethics and Dissemination:** This systematic review utilises published secondary data, thus ethics committee approval is not required. The results will be disseminated by publishing them in article format in a scientific journal.

**Trial and Registration Number:** CRD42023424225

**Article Summary:** *Strengths and Limitations of this study:*

- This is the first protocol for a systematic review designed to summarise differences in risk factors for hypertension by comparing people living with HIV (PLHIV) to uninfected people.
- To the best of our knowledge, this is the first protocol of a systematic review and meta-analysis focusing on hypertension and its risk factors in PLHIV in comparison to uninfected people that assesses the methodological quality of studies using the GRADE method;
- This systematic review imposes no restriction on language or year of publication ensuring broader coverage and more representative results;
- We will rigorously apply methodological procedures for systematic reviews to guarantee unbiased and accurate results;
- This systematic review may face challenges in identifying all potential risk factors that differ between PLHIV and uninfected people;
- The exclusion of gray literature when searching for primary studies may pose a limitation; although we assume that focusing on peer-reviewed articles will enhance the study reliability.

## INTRODUCTION

Human immunodeficiency virus (HIV) infection along with its progression to acquired immunodeficiency syndrome (AIDS), remains as one of the greatest public health challenges worldwide^1,2^. This is due to the disease’s severity, its pandemic spread, the absence of a cure, and socioeconomic barriers that hinder full access to antiretroviral therapy (ART)^3^. To date, 85.6 million people are infected with HIV globally, and 40.4 million have died due to AIDS-related diseases since the first reported cases of infection^1^.

The advent and widespread use of ART have significantly decreased the risk of infection by opportunistic diseases and neoplasms^4^. By the end of 2022, approximately 29.8 million individuals had access to treatment, contributing to a 59% reduction in new HIV infection since 1995^1^. As a consequence, PLHIV are increasingly exposed to risk factor for chronic non-communicable diseases, particularly hypertension^5^.

As a result, PLHIV are increasingly exposed to chronic non-communicable diseases risk factors, notably a rise in hypertension diagnoses ^5-7^. Research on the prevalence of hypertension in PLHIV has yielded conflicting results; some studies report an increase, while others suggest a decrease.^8,9,10^. Moreover, the specific risk factors contributing to elevate hypertension prevalence or incidence in PLHIV remain unclear^11.^ Possible influences include prolonged life expectancy leading to greater exposure to traditional hypertension risk factors, the inflammatory process induced by the virus, antiretroviral therapy use, or a combination of these factors^12-14^.

This article presents the protocol for a systematic review and meta-analysis that aims to assess the association and risk factors for hypertension in PLHIV in comparison to individuals without HIV. The development, registration, and publication of the protocol is a crucial step in a systematic review, as it ensures that all reviewers rigorously and consistently follow the predefined criteria. This not only promotes process standardization but also enhances the transparency and reproducibility of the research, minimizing biases and ensuring that the investigation is conducted in a structured, impartial, and reliable manner.

## METHODS AND ANALYSIS

### Design

This protocol was developed in accordance with the Preferred Reporting Items for Systematic Review and Meta-Analysis Protocols (PRISMA-P) guidelines^15^, as detailed in Supplementary table 1. The review is registered in PROSPERO International Prospective Register of Systematic Reviews under the registration number CRD42023424225.

### Eligibility Criteria

#### Inclusion Criteria

1. Population: PLHIV and people not living with HIV aged ≥ 18 years without geographic restriction;
2. Types of studies: Observational studies (cross-sectional, cohort, and case-control) Investigating the association between hypertension and HIV status.
3. Types of outcome: Medical diagnosis of hypertension, defined as systolic blood pressure (SBP) ≥ 140 mmHg or diastolic blood pressure (DBP) ≥ 90 mmHg^16,17^, or self-reported use of antihypertensive medication. Studies using alternative diagnostic criteria will also be included to cover the full range of available scientific literature on the subject.

#### Exclusion Criteria

1. Population: Children and adolescents under 18 years old, and pregnant women;
2. Types of studies: Case studies, case series, experimental studies, quasi-experimental studies, systematic review and meta-analysis, commentaries, editorials, conference proceedings reports, and protocols;
3. Types of outcomes: Non-systemic arterial hypertension (e.g., pulmonary, portal, and intracranial hypertension) and studies that do not specify diagnostic criteria for hypertension.

### Information sources and Search strategy

The search will be conducted across the following databases: Medline (PubMed), Embase, Web of Science, and Global Index Medicus without language or publication year restrictions. The search terms will be developed using the controlled vocabularies such as Medical Subject Heading (MeSH) and Embase Subject Headings (Emtree), synonyms, and free terms, combined with Boolean operators and truncation. The following term variations will be used for hypertension: ‘essential hypertension’, ‘isolated systolic hypertension’, ‘blood pressure’ and ‘systemic arterial hypertension’. For HIV, the following variations will be used: ‘acquired immunodeficiency syndrome’, ‘AIDS’, ‘acquired immune deficiency syndrome’, ‘human immunodeficiency virus’, ‘HIV infection’, ‘HIV’, ‘HIV patient’, ‘HIV/AIDS’, ‘HIV-positive’, ‘HIV positive patient’, ‘human immunodeficiency virus infected patient’, ‘people living with HIV’, ‘people living with HIV/AIDS’, ‘antiretroviral therapy, highly active’ and ‘highly active antiretroviral therapy’.

The primary search strategy conducted in Medline (PubMed) is outlined in Table 1. The search strategy will be adapted according to database’s specific requirements. Additionally, the reference lists of studies included in the systematic review will be examined to identify further relevant studies.

**Table 1.** Search Strategy in Medline (PubMed)

| Search | Search Terms |
| --- | --- |
| #1 | Acquired Immunodeficiency Syndrome[MeSH Terms] OR "AIDS" OR "Acquired immune deficiency syndrome" OR "human immunodeficiency virus" OR "HIV infect*" OR HIV[MeSH Terms] OR "hiv patient*" OR "HIV/AIDS" OR "HIV-positive" OR "HIV positive patient*" OR "human immunodeficiency virus infected patient" OR "people living with HIV" OR "people living with HIV/AIDS" OR "antiretroviral Therapy, Highly Active"[MeSH Terms] OR "highly active antiretroviral therapy" |
| #2 | "hyperten* "[MeSH Terms] OR "essential hyperten*"[MeSH Terms] OR "isolated systolic hyperten*" OR "blood pressure" OR "systemic arterial hyperten*" |
| #3 | # 1 AND # 2 |

### Screening of Studies

Studies retrieved from the databases will imported into the Covidence application, where duplicates will be automatically removed, studies selected and data extracted. The selection of studies will occur in three phases. In the first phase, the authors will conduct a pilot test using the first 500 references retrieved from the PubMed/Medline database to ensure that relevant studies were correctly identified and that reviewers reached a consensus on the eligibility criteria. Additionally, data from this test served as the basis for developing the data extraction form. In the second phase, three reviewers (MGG, KSC, ECP) will independently review titles and abstracts. In the third phase, the same three reviewers will independently assess the full texts of potentially eligible studies. If discrepancies arise during the second or third phase, the lead reviewer will contact the other reviewers individually to determine the reasons for the disagreements. If necessary, adjustments to the protocol will be made to resolve the conflicts before the actual systematic review starts.

### Data Extraction

Data will be extracted independently using the Covidence framework by three reviewers (MGG, KSC, ECP). Configuration for data extraction will be based on a standard form tested by the team in the pilot study. Any disagreements will be resolved by the lead reviewer (Box 1).

**Box 1.** Variables to be extracted.

| Category | Variables |
| --- | --- |
| Article details | First author's name and year of publication |
| Study characteristics | Continent, including Africa, North America, South America, Central America, Antarctica, Europe, Oceania, Asia; country; study design, including cross-sectional, cohort, case-control; data collection time, considering prospective or retrospective; follow-up period as years of follow-up; data classification as primary or secondary data; and source of secondary data. |
| Participant characteristics | Number of participants living with HIV and uninfected participants, proportion of men and women, age threshold for inclusion, and age presented as mean or median and standard deviation (SD), interquartile range (IQR), or range. |
| HIV characteristics | Time since infection presented as years, proportion of infected participants on treatment, duration of treatment presented as years |
| Outcome characteristics | Diagnostic criteria, number or proportion of hypertensive participants in both groups, systolic and diastolic blood pressure in both groups presented as mean or median and standard deviation, interquartile range, or range |
| Other Information | Body mass index presented as mean and standard deviation, body mass index category presented as absolute value, number of participants in both groups with type II diabetes mellitus, who smoke, consume alcohol, and engage in physical activity. |

#### RISK FACTORS

For the extraction of data specific to the risk factors, the following criteria will be applied to select studies:

##### Sex

Studies must categorise participants by sex (male and female) to distinguish the number of individuals with hypertension among PLHIV and those without HIV.

##### Body Mass Index (BMI)

Studies should follow the World Health Organisation (WHO)^18,19^ guidelines for BMI. The inclusion criteria will focus on studies reporting the total number of participants classified as overweight or obese, both in PLHIV and non-HIV groups, as well as the number of individuals with hypertension in both groups who are overweight or obese.

##### Type II Diabetes

Eligibility requires a physician’s diagnosis of type II diabetes based on any of the following biochemical tests: oral glucose tolerance test, fasting glucose, glycated haemoglobin, or random glucose test, as well as self-reported diabetes ^20-22^. Studies should report the total number of participants with type II diabetes in both PLHIV and non-HIV groups, as well as the number of overweight or obese participants with hypertension in both groups.

##### Alcohol Consumption

Studies using the Alcohol Use Disorders Identification Test (AUDIT) to assess excessive alcohol consumption and hypertension in PLHIV compared to non-HIV participants will be included^23^. These studies should report the number of participants with excessive alcohol consumption or binge drinking behaviour and hypertension in both groups. Excessive alcohol consumption is defined by an AUDIT score of 8 or higher, and binge drinking is classified as the consumption of five or more drinks in one session^24^.

##### Smoking

Eligible studies must classify participants as: (I) Current Smokers – adults who have smoked at least 100 cigarettes in their lifetime and continue to smoke. (II) Former Smokers – adults who have smoked at least 100 cigarettes in their lifetime but quit before the interview^25,26^. Studies should report the total number of current and former smokers, both in PLHIV and uninfected groups, as well as the number of individuals with hypertension in both groups.

##### Physical Activity

Studies must use the short version of the International Physical Activity Questionnaire (IPAQ), as recommended by the World Health Organisation (WHO), to assess physical activity levels. Sedentary participants are defined as those who did not engage in at least 10 continuous minutes of physical activity during the week^27,28^. Studies should report the total number of sedentary participants in both the PLHIV and non-HIV groups, as well as the number of sedentary participants with hypertension in both groups.

### Risk of Bias in Individual Studies

The methodological quality of the studies included in the review will be independently assessed by four reviewers (Iniciais) using the Newcastle-Ottawa Scale (NOS) for cohort and case-control studies^29^. Studies will be rated from zero to nine stars: scores of 8 or higher will indicate high-quality, scores between 6 and 7 moderate quality, and scores of 4 or lower poor methodological quality^30^. For cross-sectional studies, the Joanna Briggs Institute (JBI) Checklist for Analytical Cross-Sectional Studies will be applied^31^. The final JBI score will be based on the percentage of “yes” answers: studies with 70% or higher “yes” responses will be classified as high quality, those with 50% to 69% as moderate quality, and studies with 0 to 49% as low quality ^32^.

### Data Synthesis

The information extracted from each study will be systematically described, analysed and summarised to meet the objective of the systematic review. A meta-analysis will be conducted using a random-effects model with the Restricted Maximum Likelihood method as the variance estimator. Heterogeneity will be assessed using Cochran’s Q test, with a p-value of < 0.1 considered indicative of heterogeneity, and Higgins’ I^2^ index. The following sensitivity analysis methods will be employed: the leave-one-out technique and the removal of studies with low or moderate methodological quality. Publication bias will be examined through visual inspection of the funnel plot, and if 10 or more studies are included in the meta-analysis, Egger’s test will be applied. Additionally, a meta-regression will be performed to assess the influence of moderator variables on heterogeneity.^33^ In case a meta-analysis is not feasible, a narrative synthesis of the results will be provided^34^. The analyses will be performed using the meta package (version 7.0-0) in R studio.

### Confidence in Cumulative Evidence

The certainty of scientific evidence will assessed using the Grading of Recommendations Assessment, Development and Evaluation (GRADE). GRADE will be applied to assess each research outcome. A summary table of results will be generated using the GRADE online software (GRADEpro GTD, Copenhagen, Denmark)^35^.

## CONCLUSION

This protocol provides the methodological framework for a systematic review and meta-analysis. The study will assess the association between systemic arterial hypertension in people living with HIV (PLHIV) and in HIV-uninfected individuals. Following this protocol ensures that the research process will be rigorous, unbiased, and reproducible. This research will employ rigorous methods, following all steps guided in the literature on conducting systematic reviews. Consequently, it will enables robust synthesis of knowledge regarding hypertension in PLHIV and inform the formulation of public policies. Additionally, this research will help identify gaps in the global understanding of hypertension in PLHIV.

### Reporting of this Review and Dissemination

The systematic review and meta-analysis outlined in this protocol will be conducted following the Preferred Reporting Items for Systematic reviews and Meta-Analyses (PRISMA)^36^. The results of this systematic review will be published in a peer-reviewed scientific journal and at scientific conferences and congresses.

## Data Availability

All data produced in the present work are contained in the manuscript

## Potential Amendments

Changes made during the pilot study or the execution of the systematic review stages will be reported transparently in PROSPERO.

## Ethics Consideration

This study uses secondary data and, therefore, ethics committee approval is not necessary.

## Author Contributions

MGG and SLR were responsible for developing the protocol and both approved the final version of the submitted manuscript.

